# Trying to Heal Without “Bringing Problems”: Navigating Cervical Precancer Stigma in a Phase I Clinical Trial in Western Kenya

**DOI:** 10.1101/2025.10.24.25338728

**Authors:** Anagha Guliam, Annum Sadana, Elizabeth Opiyo, Felix Adundo, Katherine Sorgi, Erum Agha, Chemtai Mungo

## Abstract

**Background:** Cervical cancer remains a leading cause of cancer-related deaths among women in low- and middle-income countries, particularly in sub-Saharan Africa. Sociocultural barriers including stigma significantly influence women’s decisions around screening and treatment.

**Objective:** To explore how stigma influences women’s experiences with cervical precancer care in the context of emerging self-administered topical therapies.

**Design:** A qualitative approach using convenience sampling.

**Methods:** Seventeen in-depth interviews were conducted between 2024 and 2025 in Kisumu, Kenya, with women who had been diagnosed with cervical precancer. Participants were recruited to self-administer intravaginal artesunate in a Phase I clinical trial, and interviews explored their experiences. Thematic analysis was conducted using the Health Stigma and Discrimination Framework.

**Results:** All women discussed experiences indicative of enacted, anticipated, and internalized stigmas related to partner blame, disclosure, and imagining the diagnosis as an imminent death sentence. These stigmas were driven and facilitated by poverty and limited health care access, embarrassment around sexual health, community fears of cervical cancer death, patriarchal norms, and cervical precancer’s disproportionate burden on women. Women with cervical precancer feel “marked” by medication usage, clinic visits, community expectations of a “cancer” appearance, or painful sexual intercourse. By exercising relational autonomy, women in this trial were able to navigate stigmas around sexual health and overcome the debilitating fear of death. They were ultimately able to complete the self-administered artesunate treatment course and initiate conversations in their communities to address stigmas around cervical precancer treatment. Nevertheless, stigmas caused delays to screening and treatment among some participants, while others experienced intimate partner violence or conflict during artesunate treatment.

**Conclusion:** Stigmas around cervical precancer, artesunate usage, and abstinence requirements can deeply impact women’s social experiences, intimate partnerships, and mental health. Future trials of self-administered therapies should integrate stigma reduction strategies developed in collaboration with participants and survivors.

ClinicalTrials.gov NCT06165614, https://clinicaltrials.gov/study/NCT06165614

## 1. Introduction

Cervical cancer is the fourth leading cause of cancer deaths among women globally, with 662,301 new cases and 348,874 deaths in 2022 (1,2). Low- and middle-income countries (LMICs) bear a disproportionate burden of cervical cancer mortality, as 94% of deaths occurred in these countries (1,2). Sub-Saharan Africa has the highest cervical cancer mortality rate, exceeding 16 per 100,000 women-years (1). In Eastern Africa, the incidence of cervical cancer is 40.4 per 100,000 women-years, far above the World Health Organization (WHO)’s 4 per 100,000 women-years elimination target (2). To meet the WHO’s 90-70-90 target for the elimination of cervical cancer—that 90% of girls should be fully vaccinated by age 15, 70% of women should be screened for cervical precancer or human papillomavirus (HPV), and 90% of those diagnosed with cervical precancer or cancer should receive appropriate treatment—it is essential to address the various barriers to treatment in LMICs (3). These include financial inaccessibility, oncology workforce, treatment-related fears, concerns about side effects, lack of knowledge, and sociocultural factors (4–8).

A significant sociocultural barrier to cervical cancer screening and treatment is stigma, particularly due to dual stigmas of a potential cancer diagnosis and infection with human papillomavirus (HPV), a sexually transmitted infection (4,7,9–12). Stigma is the process by which individuals are identified as socially undesirable and therefore marginalized due to attributes or behaviors that are considered “deeply discrediting” (13,14). Those in power may subject stigmatized individuals to labels, stereotypes, separation, status loss, and discrimination based on a health condition (15).

In an LMIC where there are many barriers to screening and treatment, understanding and addressing stigma is crucial to improving cervical cancer prevention and care. Previous studies have examined stigmas associated with cervical cancer screening and provider-administered treatment. A recent qualitative study with 26 in-depth interviews in Kisumu, Kenya previously used this framework to describe how community knowledge and misinformation around HPV risk factors, cultural beliefs, and cancer fatalism could drive stigma. Stigma manifested as discriminatory attitudes, anticipated stigmas around gossip, partner support, and medical mistreatment, and internalized stigmas such as hopelessness and embarrassment. Collectively, these led to a decreased perception of risk, and reduced cervical cancer screening and treatment uptake (11). Cervical cancer screening may involve stigmas around HPV infections, as women may be blamed for being promiscuous, a notion perpetuated by pervasive male chauvinism (6,7,16). In several qualitative studies, women felt it was necessary to hide an HPV infection, as it may be perceived as shameful and embarrassing (6,7,12,16,17). They also feared spousal and societal disapproval of seeking pelvic exams or treatment (6,7,12,16,17). In a study conducted in Nairobi and the Nyanza region of Kenya, women feared marital discord because of a cervical cancer diagnosis, especially if they could not bear children (7). Male partners admitted that others in their communities might be less understanding or even violent if women pursued treatment, assuming infidelity or reacting negatively to abstinence requirements (7). In our previous study on acceptability of self-administered topical therapy for HPV or cervical precancer in Kenya, nearly all men (99%) interviewed were willing to abstain from sex and supported women using self-administered treatment (18). However, men said that their partners might fear asking them to use condoms during topical precancer treatment due to concerns about potential violence (18). Women also feel that they need to “negotiate sexual abstinence” in a patriarchal system where they have limited abilities to make their own health decisions, such as when accepting HPV, cervical precancer, or cancer treatment options that require abstinence (19). Fatalism, denial, avoidance of a positive diagnosis, and fear of social isolation due to cancer stigma pose barriers to screening and treatment uptake (4,6,11,16,17). Women living with HIV (WLWH) in particular, face an “intersectional stigma,” as the possibility of a cervical cancer diagnosis elicits a fear of exacerbating their existing “health burdens” (11).

Medical interventions must address the various forms of stigma, as they can affect health behaviors, mental health outcomes, and interpersonal relationships that can in turn impact medication adherence (14). To achieve the WHO goal of treating 90% of women with cervical precancer by 2030, self-administered topical therapies currently under investigation may overcome key barriers to ablative and excisional cervical precancer treatments in LMICs, namely limited access, painful procedures, high costs, and lack of privacy during pelvic examinations (NCT05413811) (9,20). In our previous studies on the acceptability of a potential topical intravaginal treatment such as 5-fluorouracil (5FU) and artesunate in Kenya, most women and their partners preferred self-administered treatment at home over provider-administered therapies (18,21,22). Yet, in qualitative interviews, finding a private place to self-administer, condom use during 5FU treatment, abstinence requirements, and disclosure to partners continued to be sources of concern (9,18,21,22). Self-administered treatments, while promising, may be impacted by sociocultural norms that can hinder their use. Therefore, it is important to understand how stigmas surrounding HPV, cervical precancer, and self-administered topical therapies affect a woman’s experience and well-being throughout her treatment journey, as well as how she navigates these challenges. The Health Stigma and Discrimination Framework (HSDF) is an ideal way to map how health stigma drivers, facilitators, and manifestations operate across socio-ecological levels (23). A feminist lens of “relational autonomy” illuminates how women’s health decisions are shaped by interpersonal relationships and broader structural contexts. It also shows how women exercise agency through the support and care embedded in their social networks (24).

Little is known about how stigma impacts participants undergoing self-administered treatment for HPV and cervical precancer in an LMIC. In this manuscript, we draw from the HSDF and a relational autonomy lens to describe how women navigate stigma while self-administering intravaginal artesunate in Phase I clinical trial in Western Kenya.

## 2. Methods

### 2.1 Study setting, design, and recruitment

This study was conducted at the Lumumba Sub-County Hospital in Kisumu County, Kenya from March 2024 to May 2025. Although located in an urban area, the hospital provides HIV care to patients across Western Kenya, with a catchment area of approximately 3 million, from Kisumu, Busia, Homabay, and Vihiga counties, most of whom live in in peri-urban or rural settings. Study participants were referred from cervical cancer screening sites across the region.

In our Phase I pilot clinical trial (ClinicalTrials.gov NCT06165614), participants with histologically confirmed cervical intraepithelial neoplasia 2 or 3 (CIN2/3) were recruited to self-administer a 200 mg artesunate pessary nightly, 5 days a week (25). The 8-week regimen consisted of alternating dosing and rest weeks, with clinic visits scheduled during rest weeks.

Detailed inclusion and exclusion criteria, study procedures and recruitment methods have been previously reported (26).

### 2.2 Research team

The principal investigator, CM, is a Kenyan-born practicing obstetrician/gynecologist. She developed the interview guide, building on prior qualitative work in Western Kenya. Interviews and translations were conducted by EO, a Kenya-based clinical researcher with over a decade of experience in HIV, mental health, and stigma reduction interventions in Kisumu County. Her professional and research background in counseling, interpersonal psychotherapy, and community-based HIV care, together with her perspective as a Kenyan woman, informs her understanding of the intersections of mental health and HPV-related stigma. AG and AS are first-generation medical students with backgrounds in medical anthropology and social policy, respectively; they developed the codebook and conducted thematic analysis with the support of EA, who has extensive experience in qualitative sociocultural research and social work. They began data analysis by engaging in reflexivity discussions and examining their positionalities and potential biases as women and U.S.-based researchers undergoing medical training.

### 2.3 Data collection and analysis

As part of the clinical trial, in-depth interviews (IDIs) were conducted at the end of the dosing phase, focusing on the acceptability and challenges of self-administered artesunate, partner support, and community perceptions of cervical precancer and treatment. Seventeen participants completed the hour-long IDIs in their preferred language, Luo or Swahili, in a private room in the clinic. Following each IDI, the interviewer, EO, maintained audio recordings of each interview and a written memo about participant body language and other observations.

She also transcribed and translated IDIs to English. Through an iterative consensus-coding process, EA, AG, and AS developed a codebook using Dedoose 10.025. Data saturation was reached at 15 IDIs. The topic of stigma emerged inductively, and we did thematic analysis guided by the Health Stigma and Discrimination Framework (HSDF) and relational autonomy lenses (23,24).

### 2.4 Conceptual frameworks

The HSDF is a theoretical model to describe the intersection of health stigma with stigmas related to race, gender, sexual orientation, class, and occupation. It includes drivers, which are “inherently negative” community-level forces that cause stigma, and facilitators, which can “exacerbate stigmatizing” behaviors. Stigma markers are traits which label certain individuals or groups. Stigma manifestations can be experienced, anticipated, or internalized. Outcomes of these stigma manifestations may include the (in)ability to access healthcare services, adherence to a medication, quality of health services, law enforcement practices, and social protections (23). We draw from the HSDF to outline our themes around drivers, facilitators, manifestations, and outcomes of cervical precancer stigma during self-administered artesunate treatment.

We also use a relational autonomy lens to interpret the interpersonal and structural factors that influenced participants’ experiences maneuvering through complex social relationships and stigmas. Our analysis is guided by Bell (2020)’s recommendations for applying the feminist paradigm of relational autonomy to qualitative health research (24). Classical views of autonomy tend to be individualistic, rationalistic, and masculinist, as they “isolate the patient as decision maker” and ignore the social relationships that often influence a woman’s agency (27,28). In contrast, relational autonomy is grounded in the fact that each person is shaped by their social relationships and intersecting social identities (such as gender, income level, race, and ethnicity). A relational autonomy lens allows us to observe how social relationships “impede or enhance” patients’ capacities for autonomy (28). Through this lens, we identified how women in Kenya navigated stigma within their social environments while self-administering intravaginal artesunate.

### 2.5 Ethical clearance

This study was approved by the ethics review boards at University of North Carolina Chapel Hill and Amref ESRC Kenya. Written consent for the IDIs was obtained by the study clinical team, and participants were informed that the research team’s primary goal was to better understand their experiences using artesunate. Participant confidentiality was ensured through de-identification of all excerpts by assigning each IDI number with a corresponding randomized letter.

## 3. Findings

### 3.1 Demographics

Seventeen participants completed IDIs. Their mean age was 39.9 years (SD = 6.82). The majority had a primary level of education or less (64.7%), and one participant had completed secondary education. Most participants (82%) reported earning less than 500 Kenyan Shillings (KSH) per day (approximately USD 4). They predominantly worked in business, trading, or vending (58.8%), followed by farming (11.8%), salaried employment (5.9%), and other professions. The majority of participants were women living with HIV (88.2%), most of whom had been diagnosed more than 10 years ago (73.3%). Of the 17 participants, 10 (58.8%) reported having a current partner.

### 3.2 Health Stigmas

**Figure 1.** Perceived facilitators, drivers, markers, manifestations, and outcomes of stigma, and the role of relational autonomy in mitigating its impact among women receiving self-administered artesunate treatment for cervical precancer in Kisumu, Kenya.

### 3.3 Stigma Facilitators and Drivers

Adapting the HSDF, we summarize our themes in Fig 1. Factors that contribute to stigma are either stigma “facilitators” or “drivers.” Here, we refer to facilitators as enablers of stigma and drivers as more direct causes of stigma (23). Facilitators and drivers that emerged from our interviews are outlined in Table 1.

Poverty and a limited access to health care emerged as powerful facilitators of cervical precancer stigma, as unaffordable treatments led to worsening symptoms, invasive procedures, and cervical cancer deaths. Seeing late-stage cervical cancers marked by visible pain, weight loss, and disability intensified community fears of a cervical precancer diagnosis. In turn, many women felt discouraged from seeking screening, and conversations around cervical precancer and were halted by anxiety and avoidance. Embarrassment around pelvic exams and anticipated pain during pelvic exams facilitated stigmatizing behavior. Even when participants attempted to normalize screening, women in their communities expressed strong discomfort and discouraged open conversation. Meanwhile, patriarchal gender norms drove men’s stigmatizing behaviors as they contributed to men feeling entitled to sex at any time as their conjugal right. If women did not fulfill these expectations, men could, at times, respond with violence or extramarital affairs. Meanwhile, the social framing of sexual health topics as taboo or “dirty” caused discomfort around discussing them. Participants shared that this may hinder the disclosure of sensitive issues like intimate partner violence (IPV) or sexual coercion to health care providers. Furthermore, women felt strongly that they bore a disproportionate burden of HPV-related diseases and the stigma of cervical precancer. Women were expected to seek treatment and heal quickly to carry out their duties as breadwinners and mothers. This added an enormous burden and great expectation for them to carry, leaving women vulnerable to bearing the weight of cervical precancer stigmas (Table 1).

### 3.3 Stigma Markers

Stigma drivers and facilitators exploit identifiable markers of cervical precancer to devalue patients (23). The following markers emerged in our interviews: (1) medication usage and storage, (2) clinic visits, (3) painful sexual intercourse, and (4) community expectations of a ‘cancer appearance’ (Table 2).

### 3.4 Manifestations of Stigma

#### Anticipated Stigma

Anticipated stigma occurs when expecting or imagining future discrimination based on an illness (23). Partnered and unpartnered women alike anticipated IPV and partner rejection upon disclosure of their cervical precancer diagnosis and treatment. One participant’s brother ended his marriage after his wife tested positive for HPV, assuming that she was having an affair (IDI F). The participant subsequently felt that the same could happen to her.

> Challenges are many because most men are different like maybe you have a drunkard partner, mine is also not supportive and do not care how you do your things and sometimes he may think that you are having extra marital affairs or you don’t want him in your life and for most men to understand that you are on medication it will take some time. (IDI F)

An unpartnered participant anticipated that women could face sexual coercion or physical violence if they deny sex after inserting artesunate and the tampon.

> *Like sometimes someone’s partner is alcoholic, and he does not understand that you are on treatment inserting the drug. Like he does not care whether you are using the drug or not and sometimes he wants to have sex with you whether you like it or not, but sometimes to avoid the beatings you will have to remove the tampon and drug is already inside working. (IDI K)*

The anticipated stigma of IPV may cause women to face conflict, delay disclosure, remove the tampon shortly after insertion, or skip using the medication.

> *When I Initially started, the side effect that I almost experienced is that I did not tell my husband that when you have inserted the medicine, we cannot have sex, but I later sat him down and explained to him and he told me that I should have told him earlier. (IDI M)*

> *Some women do not tell their partners what they are using and when they want sex, they will skip using the medication due to fear. (IDI O)*

Participants also feared disclosing their condition to others in the community due to anticipated gossip about their diagnosis and intimate relationships, as well as the potential for discouragement during treatment.

> *Maybe, he shares it with your mother-in-law and other in-laws and when you are around, they are quiet not knowing what is going on because most men are not secretive. Most of the time it is us women who keep a lot of secrets and even if you go to the hospital and they test positive most women will endure a lot, but for men they will tell everyone about it and the whole family will know your status. From the mother-in-law telling your other co-wives and the circle goes. It is always not an easy issue. (IDI F) It is something that you use at night because there are some people who will enquire more to know the drug you are taking; others will discourage you that you are taking cancer drugs. (IDI J)*

#### Enacted Stigma

Stigma was enacted upon women through discrimination and prejudice related to their precancer diagnosis and treatment. This manifested when misinformation about HPV infection and precancer treatment contributed to community beliefs that healing would not be possible if a woman continued to have sex during treatment. One participant faced stigma for taking medication without her partner. This not only implied a misunderstanding of HPV transmission but also raised suspicions about her relationship.

> *When I was going for treatment, I was always told that I should go with my partner and when they give me drugs, he should get his drugs to take, they were asking why I was taking the drugs…because as I take the drugs and we continue having sex I will not heal because of the recurring infection. (IDI I)*

Some partners were not understanding of the treatment process. One of the participants described a partner who accused hospital staff of causing conflict in their marriage. Another participant was emotionally abused by her partner. She expressed concern for those who may face physical or sexual violence when undergoing self-administered artesunate treatment.

> *…he will tell you that the hospital staff are the people who are causing that conflict. So, he will tell you that since you started going to the hospital for treatment you are tough headed, and you are not but he does not know that you are sick because you are doing all that to get healed and he does not understand. (IDI D)*

*That day I did not sleep at my home. I was from the market, I had taken a shower and inserted the medicine. Then he told me, “Women face this side.” Because he could not beat me, being that he is paralyzed. I didn’t want to have a confrontation with him, because he told me to leave his bed. So, I was feeling like, this man is sick, what of someone else who just has a healthy husband? Don’t you think they can just forcefully have sex with their wives when they have inserted the medicine? I just had such a thought in my mind. That women can face such challenges. (IDI L)*

Women also encountered enacted stigmas when community members commented that they would not heal from cervical precancer, even after treatment with artesunate. For example, a participant shared, “The local herbalist who feels that this medicine is not going to heal me, has told me…that they don’t treat such conditions in the hospital” (IDI H). Some community members told harrowing stories of cervical cancer death, insinuating that the same may be true for the participants.

> *The lady who came to my house at that time and told me that it is cancer, saying that is how cancer affects women and was telling me that it affected another woman and she died and another one also went to the hospital and never came back…She was telling me that cancer spreads so fast and I did not answer her because all the questions and what people were talking about with a lot questions hurts more. (IDI N)*

#### Internalized Stigma

Internalized stigma occurs when an individual accepts social devaluation and negative beliefs related to their stigmatized condition (23). For many participants, this manifested as an anxiety around cervical precancer being equivalent to a death sentence. Poverty further exacerbated this anxiety, as women felt hopeless that most treatments involved a significant cost barrier.

> *Most of the time you will find that majority of the people fear the word cancer, and from the call I received and my knowledge that I should start treatment immediately I just knew it is cancer….I knew that cancer was cancer…Most of the time you will hear news about cancer in last stages and if you get such news you just know that you are almost going for eternal rest. So that made me fear because I always test myself and what I was told that I need to start treatment immediately. (IDI J)*

> *In the year 2021, I was told that I had cancer and I used to be indoors crying all the time, so all that pains me was that I had was that I had cancer, even if I sell all household items and my home, I cannot manage the treatment and the only thing I was seeing was my death. (IDI B)*

To them, cervical precancer was a particularly “heavy” diagnosis with uncertain artesunate treatment outcomes. Several participants discussed their anxieties through physiological manifestations such as breathing fast, headaches, and sleeping longer than usual.

> …*that time I had a heart problem, and I was breathing so fast which I feel was not good when I go for surgery…I was still depressed with so much worries and while in my house I was feeling like tents was being assembled in my compound with one lady. (IDI N)*

> *So when I sleep, I always sleep well until morning, then I remove the tampon and that made me ask myself many questions about why I am sleeping, this or was it because of the drugs I was using, and they have some antidepressants inside them. (IDI B)*

Filled with anxiety around death, one participant flogged her children while another constantly imagined her funeral. Both women expressed sadness that their anxieties and depression had also affected their children. These reactions can be seen as maladaptive coping responses, reflecting the immense burden of managing their diagnosis in isolation.

> *I was angry all the time and if any child came to me, I was flogging them because hearing the word cancer it is not easy to accept it…I was acting that way because I knew cancer had no cure and my life will end like that. (IDI E) I was just thinking about death because everywhere I was what my mouth was saying is different from what I was thinking in my mind because most of the time, the picture that I had is that I died in Russia hospital and people are coming to welcome me for my final welfare before I get buried. It happened all the time and even when someone called me, I will not respond but just assume…I had a picture of how everything was going to happen like my burial and how my children’s life changed after my death. So, at that time my child came back from school and asked me my progress, and I told him that I am just resting but looking at how he was looking at me showed how depressed he was. (IDI N)*

### 3.5 Relational Autonomy: Mitigating and Coping with Stigma

Amid the various manifestations of cervical precancer stigma, participants found ways to cope and relied on their support systems to help mitigate their impact. This reflects relational autonomy, as participants’ navigation of stigma was not done in isolation, but was rather shaped by their relational contexts (24,28). For instance, the enacted and anticipated stigmas around partner rejection and IPV were primarily mitigated by communicating with partners to negotiate weeks of sexual intimacy as well as partner support of abstinence. In addition to fulfilling their partners’ intimacy needs, women used break weeks to rest from treatment and the tiring labor of being mothers and an income source for the family. Partner support, rather than blame, was important to provide time for rest.

> *You can be free to share because they always say that it is what make you be together. Like you can tell him that this week you are on treatment and the other week you will skip to enable you to engage in sexual relationship not to allow the man to complain of a lot of things like I have heard that she is on cancer treatment or any other complaints like a mad person. (IDI F) You need to rest for some time for this drug to work because if you are farming you get tired and even if you are a business person you will have worked and sometimes you can forget to apply the drug…So when it’s evening I can decide to do something or rest and my partner always tell the children that I am not feeling well and they do everything. (IDI B)*

A few women said that if their partners had acted as a barrier to the treatment process, they may have even separate from them for the sake of their own health and the wellbeing of their families.

> *Like for me, if I tell my partner about it and I do not get any support for my health, I can take a step to separate from him and seek treatment and get healed. (IDI D)*

Coping with internalized stigmas, such as anxieties, hopelessness, and the psychological burden of cervical precancer, required strong support systems. After reading information about the diagnosis, some partners provided emotional reassurance that the treatment would bring healing (IDI E). Despite stigmas around discussing sexual health topics with family members, one participant shared that she disclosed her symptoms and sexual activity to her sister, who encouraged her to seek screening (IDI H). Another participant’s mother urged her to share her diagnosis with her husband despite her fears around the diagnosis (IDI F). A son’s curiosity to learn about artesunate self-insertion encouraged a participant to openly discuss her treatment rather than hide it (IDI J). Patient education also helped dispel the misconception that cervical precancer is a death sentence, which had been fueling internalized anxiety. When participants learned about the distinction between precancer and cancer, the stages of cervical precancer, and treatment options such as artesunate, they felt relieved and confident. Surrendering to God helped women remain optimistic and resilient. While discouraging remarks about healing and internalized stigma often left them feeling exhausted, hopeless, and vulnerable, their faith provided strength throughout the many medical and social uncertainties they faced.

> *Like if you hear that you have cancer you just feel tired and if it’s not God’s grace you cannot have that strength. So even if you are in a crowd of people you feel down and with God’s grace that will make you feel good and happy pushing on that makes you feel at peace. (IDI B)*

### 3.6 Outcomes

#### Individual-level

All participants successfully completed the self-administered artesunate treatment course, including tampon use, self-insertion, and study visits despite prevailing stigmas (29). Yet, some participants discussed barriers that they faced along the treatment journey or raised concerns that other women may face.

For example, one woman faced delays in treatment due to her fear of a cancer diagnosis and her duties as a mother and financial support to her family.

> *I was very sick due to fear of cancer, stayed in the house for two weeks and I was very thin because of losing weight and my partner called my sisters and brothers to come and talk to me. After that call, they came and talked to me and the cancer diagnosis was still in my heart. So, all that was still tough and at that time we had many teenagers in high school, there were also many challenges and I had to go to Nairobi to be employed as a house manager…and that lady did not know that I have precancer and I continued doing my work until I was forgetting. So, with screening, I did it in Russia Hospital and when my first sample was taken it got lost, but I waited for too long until I went to Nairobi for two years and came back. (IDI B)*

Eventually, all the participants were able to access cervical cancer screening and treatment. However, they discussed that other women face barriers to screening and treatment due to the high cost of medical care, stigma, and fear of a cancer diagnosis. One participant compares this fear to HIV and AIDS stigma, where such a diagnosis was seen as a death sentence. She felt that with more health education and awareness of treatment options, more women may be able to overcome their fears of the diagnosis to receive necessary screening before symptoms worsen.

> *Most people fear to come out to know their status like during the HIV/AIDS era, more people were fearful to know their status and they thought that was the end of the world. So even now, more people fear to know their status and it is better you go early to know what you are going through. It is better you know early that diseases like cervical cancer are serious sicknesses and if they come you counsel them and tell them what to do. You do that for them to come for treatment because some are willing to come for treatment but they are afraid. (IDI G)*

To improve the accessibility of screening, it is not sufficient to address just a single dimension of stigma. For example, even when male partners expressed support for screening, some women still reported discomfort with pelvic examinations. These experiences highlight the multifaceted nature of stigma, which must be addressed in a comprehensive manner.

> *In my village, some doctors were walking door to door encouraging women to go for cancer screening and some male partners are encouraging their female partners to go for screening and they are afraid to go for it saying they cannot open their legs for something to be inserted in their vagina they will not accept that. (IDI E)*

The same social and gendered stigmas that inhibit screening also complicate treatment. Concerns about abstinence, partner control, and sexual violence emerged as barriers to proper use of artesunate. While only one participant reported a violent incident that led to a missed dose, others feared that coerced sex from a partner could interfere with self-insertion or adherence to treatment protocols.

> *Remember some of us do not have refrigerators in our houses and I cannot put the medicine in my pocket, I must just put it in a cold place and sometimes, and sometimes I place it under the pot when he becomes violent and I run, before picking my medicine, it will force me to miss using the medicine for that day, and there is no way for me to go pick my medicine. One will end up not using the treatment as required. (IDI L)*

> *The major challenge is on men who are not consistent and they just come at any time when you are on medication. Like women who have many sexual partners, they cannot plan and one of the male partner can come at any time and sometimes it takes long before they meet and they will have to engage in sex because I cannot say that all women are married, but at times it forces them to ignore using medication to maintain that sexual intimacy. (IDI F)*

### Addressing stigma

While completing artesunate treatment for cervical precancer, participants initiated dialogue within their communities about screening and treatment, actively working to destigmatize pelvic exams and care provided by male clinicians. They emphasized the importance of sharing their own experiences with other women: “because it can happen to anyone and so you have to be comfortable and talk, it’s when you will get encouraged” (IDI M). They desired to have an impact on their communities and saw themselves as part of the advocacy team. One participant shared, “our main agenda is to create awareness of cancer screening and then seek treatment” (IDI E). Some were successful in convincing women to get screened and referred to treatment.

> *… some women are not comfortable with male doctors screening them, especially if they lie naked to the doctor. Then I tell them there are female doctors who also do screening, they are like they still won’t feel comfortable. Then I tell them both female and male providers are just as good according to my experience, I tell them, how do they feel uncomfortable yet they are living with their husbands? And then one of them concludes that “I agree with you.” I convinced one and she told me that in the coming week she will go for screening, so that she can know her status regarding cervical screening. I then told her to go for screening so that she can give me feedback, so that I can feel how I have impact to her. (IDI M)*

In addition to engaging in conversations to encourage women to get screened, some participants also “signed for Chama money,” an informal women’s savings and support group, as a form of advocacy. This pooled money system helped cover costs associated with screening, such as transportation. Since cost was identified as the most significant barrier to accessing screening and medical care, participants felt a strong sense of responsibility to reduce financial obstacles and prevent delays to care.

> *In my village, there was one woman who approached me and had the same symptoms that I had before screening, feeling pains in the stomach and I told her to go to Emuhaya health facility for cancer screening. So for her she is always very fast and since I am the person who sign for Chama money, I signed for her and she came back that she tested positive after screening and has been transferred to Russia and the Chama money (group money) signed to be her transport yesterday and I know that today she is in Russia (IDI E)*

Despite their efforts, some participants found that women in their communities remained ashamed and fearful of screening. One participant, after advising women with abdominal pain to seek early screening and treatment, observed that they still “fear going for screening, some ashamed and uncomfortable opening their legs” (IDI B). Furthermore, the emotional burden of receiving a cervical precancer diagnosis, coupled with witnessing the barriers to care in their communities, made many women feel a personal responsibility to advocate for screening. One woman described feeling guilty when sharing the challenges of her own precancer journey, such as painful screenings, repeated pelvic exams, and exhaustion after clinic visits, with her friend.

She hesitated to speak openly about the negative aspects of her experience, as she wanted to be a strong advocate for screening and treatment. In her attempt to champion these efforts, she had inadvertently limited open conversations about the difficulties involved.

> *…the problem with us women like when I come and I leave the screening room, I will discourage another woman that screening is painful or when I talk about it, I should encourage another woman to go for cervical cancer screening and not mention my experience about it because sometimes there are women who have not gone for screening or maybe she is suffering silently and need that medical attention. (IDI K)*

In addition to facilitating dialogue around cervical precancer, women hoped to prevent delays to treatment and cancer mortality through their participation in research. By becoming research participants, they led the way to break the stigma that cervical cancer is akin to a death sentence. One participant hoped that others could “follow [her] to get treatment” (IDI E).

Another participant similarly shared that “for a person who has passed through this treatment, it is better to create awareness in the community and not keep [artesunate] for ourselves” (IDI J). Participants also hoped to encourage other women to participate in research to overcome fear and take action, rather than continuing to suffer.

> *So, after participating, you will know the progress and whether it is working or not and something must be tested first for it to work, if you use it, you will have more experience with it and testify how it helped you. As for me, I can say that you only fear when you are not sick when you go through something, that fear will not be there because if you fear and you have it without taking action you will suffer. So am encouraging women to come out. (IDI I)*

#### Participant Recommendations

Participants provided recommendations to address stigma-related barriers to cervical precancer screening and self-administered artesunate treatment (Table 3).

## 4. Discussion

Based on qualitative data from in-depth interviews with women diagnosed with cervical precancer and enrolled in a clinical trial in Western Kenya, we examined how participants navigated stigma during their treatment while exercising relational autonomy. Observing cervical cancer deaths due to poverty and limited access to timely diagnosis and treatment fueled community doubts about recovery. Comments from others that participants would not heal even with treatment heightened women’s anxieties and sometimes delayed care. This is in line with literature linking financial barriers to delayed treatment and mortality (4,5,10) as well as qualitative studies showing that fear of cervical cancer death can discourage screening and treatment uptake among screening-eligible Kenyan women (7,11,17). Women drew on partners, family, peers, and clinical teams for emotional support to cope with internalized stigma, echoing qualitative findings that peer and partner support encourage screening and clinical trial participation (17,30,31). They also expressed a desire for greater partner involvement during treatment, consistent with results from a qualitative study on same-day treatment with thermocoagulation in Malawi (19). At the same time, patriarchal norms, stigma around sexual health during pelvic exams, and family demands intensified women’s burdens, as seen in other LMIC contexts (5,6,11,32). Anticipated and enacted stigma related to abstinence, relationship strain, and partners’ limited understanding of HPV align with findings from systematic reviews and regional studies that explored participants’ perspectives on screening, HPV-positive diagnoses, partner disclosure, and hypothetical cervical precancer treatment (6,7,9,12,19,33).

Many of our participants were WLWH, and some drew comparisons between artesunate and ARVs and noted similar fears that could act as barriers to care. Prior research has shown that WLWH may perceive less stigma around cervical cancer than HIV-negative women, possibly because they have already undergone a destigmatizing process (34).

### Strengths

Prior studies have examined cervical cancer stigma mainly as a barrier to screening and, to some extent, provider-administered care. To our knowledge, ours is the first to analyze how participants navigated stigma during a trial of self-administered intravaginal cervical precancer treatment in an LMIC. Participants discussed a spectrum of challenges and stigmas faced, from noticing symptoms and overcoming barriers to screening, to deciding to participate in a clinical trial and managing self-administered artesunate. Furthermore, women in this study became advocates for screening and treatment by signing for chama (group) funds, spreading hope and awareness about treatment options, and initiating dialogue to destigmatize pelvic exams and encourage partner engagement.

### Limitations

The in-depth interview questions focused on the acceptability of self-administering intravaginal artesunate and did not explicitly address stigma. Therefore, the present study represents a secondary analysis of these interviews with a new focus on stigma. Additionally, our study cannot be generalized to stigmas and outcomes of all women diagnosed with cervical precancer. Participants were recruited at the point of diagnosis, so women most affected by stigma may have opted out of participation in screening. This means that stigma may not have been as significant a barrier for our sample as it is for others in the broader community.

### Study Recommendations

Self-administered treatments like artesunate may reduce some markers of stigma, such as fewer clinic visits or using the treatment discreetly at night in a private location, but they do not eliminate stigma entirely. Women can still be ‘marked’ by key aspects of self-administered topical therapies, including the need for sexual abstinence, possible suspicion when a woman has to hide the medication in a neighbor’s house due to lack of privacy at home or fear of disclosure, and the challenge of using the medication when partners or family members are present.

Therefore, it is important to address all levels of stigma across the HSDF. Culturally informed counseling for participants is one way to address internalized stigmas, including anxieties around death, hopelessness, and depression (17). Mental health, poverty, and stigma are often interconnected. These can place a financial and emotional burden on families, who may also need support (35). Studies focused on cervical precancer treatment or other sexual and reproductive health topics should also incorporate questionnaires on IPV. Future studies could administer patient-centered psychosocial assessments before starting treatment to lay the groundwork for a relational autonomy-based approach. These assessments may ask about key relationship ties with others, roles the patient wishes others to play, and other contexts of a patient’s life and experiences (27) to inform targeted counselling and support during clinical trials.

Participant efforts to destigmatize cervical precancer treatments can be strengthened through training and involvement in community-based awareness programs. For example, a teaching hospital in Kenya implemented a multicomponent navigation strategy involving lay navigators, clinical officers, and survivors, which helped patients complete chemotherapy-related tasks. Video-based education with motivational survivor stories, combined with assistance for health insurance enrollment and stipends, addressed both stigma and cost barriers. These interventions increased knowledge and awareness, fostered a sense of belonging, and instilled hope for survival among people living with HIV and Kaposi’s sarcoma (36). Similarly, an ongoing study, Elimisha HPV (ClinicalTrials.gov NCT05736588) (37) aims to provide clear messaging around HPV and the benefits of screening through videos, peer perspectives, and peer navigation that will be integrated with HIV-care clinics in Western Kenya.

Finally, it is essential to address two key drivers of cervical precancer stigma—poverty and cost barriers that perpetuate cervical cancer mortality. Researchers should continue developing and implementing low-cost diagnostic and therapeutic approaches appropriate for local clinic settings, while also providing travel stipends and other financial assistance as needed (5,8,36). Just as expanded access to antiretroviral therapy helped reduce HIV stigma (38), increasing access to treatments like self-administered artesunate, alongside greater public awareness, holds promise for reducing HPV and cervical precancer stigma.

## 5. Conclusion

This study applies the Health Stigma and Discrimination Framework to understand stigma experienced by women undergoing self-administered artesunate treatment for cervical precancer in Western Kenya. By outlining the stigmatization process and the protective role of relational autonomy, we identify opportunities to support women in coping with internalized cervical precancer anxieties, fostering partner and peer support, addressing intimate partner violence, and strengthening health education and community outreach to dispel misinformation. Future research on stigma as it relates to cervical precancer and HPV should focus on how stigma may influence the abstinence requirements following existing cervical precancer treatments such as thermal ablation and LEEP and further investigate any stigmas specific to intravaginal treatment under study.

## Data Availability

All data produced in the present study are available upon reasonable request to the authors.

## List of abbreviations

LMICs: Low- and middle-income countries:
WHO: World Health Organization
CIN2/3: High-grade cervical intraepithelial neoplasia
HPV: Human papillomavirus
WLWH: Women living with HIV
ARV: Antiretroviral
IPV: Intimate partner violence
HSDF: Health Stigma and Discrimination Framework

## Acknowledgements

The authors are grateful to the Phase 1 study participants for openly sharing their experiences with stigma and intravaginal artesunate. We thank the research teams at KEMRI for their support at the study site, and in particular, Lizanne Orao for her review of transcripts, feedback, and provision of cultural context during thematic analysis. We also acknowledge Konyin Adewumi for her assistance with coding IDIs and for contributing her qualitative research expertise.

Artesunate vaginal suppositories (pessaries) were provided as in-kind support by Frantz Viral Therapeutics (Mentor, OH).

## Authors’ contributions

AG: Data curation, formal analysis, visualization, writing of the original draft, writing – review and editing. AS: data curation, formal analysis, writing –review and editing. EA – analysis and supervision, writing – review and editing. EAO conducted, transcribed, and translated interviews. FA contributed to data curation and analysis. KS assisted with project administration. CM conceived of the trial, qualitative interview questionnaire and contributed to writing – review & editing.

## Statements and Declarations

### Ethics approval and consent to participate

This study has full ethics review board approval from the University of North Carolina Chapel Hill and Amref ESRC Kenya. Written informed consent was obtained from all study participants.

## Consent for publication

Not applicable.

## Availability of data and materials

Not applicable.

